# Viral RNA load in plasma is associated with critical illness and a dysregulated host response in COVID-19

**DOI:** 10.1101/2020.08.25.20154252

**Authors:** Jesús F Bermejo-Martin, Milagros González-Rivera, Raquel Almansa, Dariela Micheloud, Ana P. Tedim, Marta Domínguez-Gil, Salvador Resino, Marta Martín-Fernández, Pablo Ryan Murua, Felipe Pérez-García, Luis Tamayo, Raúl Lopez-Izquierdo, Elena Bustamante, César Aldecoa, José Manuel Gómez, Jesús Rico-Feijoo, Antonio Orduña, Raúl Méndez, Isabel Fernández Natal, Gregoria Megías, Montserrat González-Estecha, Demetrio Carriedo, Cristina Doncel, Noelia Jorge, Alicia Ortega, Amanda de la Fuente, Félix del Campo, José Antonio Fernández-Ratero, Wysali Trapiello, Paula González-Jiménez, Guadalupe Ruiz, Alyson A. Kelvin, Ali Toloue Ostadgavahi, Ruth Oneizat, Luz María Ruiz, Iria Miguéns, Esther Gargallo, Ioana Muñoz, Sara Pelegrin, Silvia Martín, Pablo García Olivares, Jamil Antonio Cedeño, Tomás Ruiz Albi, Carolina Puertas, Jose Ángel Berezo, Gloria Renedo, Rubén Herrán, Juan Bustamante-Munguira, Pedro Enríquez, Ramón Cicuendez, Jesús Blanco, Jesica Abadia, Julia Gómez Barquero, Nuria Mamolar, Natalia Blanca-López, Luis Jorge Valdivia, Belén Fernández Caso, María Ángeles Mantecón, Anna Motos, Laia Fernandez-Barat, Ricard Ferrer, Ferrán Barbé, Antoni Torres, Rosario Menéndez, José María Eiros, David J Kelvin

**Affiliations:** Group for Biomedical Research in Sepsis (BioSepsis). Instituto de Investigación Biomédica de Salamanca, (IBSAL), Paseo de San Vicente, 58-182, 37007 Salamanca, Spain; Hospital Universitario Río Hortega, Calle Dulzaina, 2, 47012 Valladolid, Spain; Centro de Investigación Biomédica en Red en Enfermedades Respiratorias (CIBERES), Instituto de Salud Carlos III, Av. de Monforte de Lemos, 3-5, 28029 Madrid, Spain; Department of Laboratory Medicine, Hospital General Universitario Gregorio Marañón, Calle del Dr. Esquerdo, 46, 28007 Madrid, Spain; Department of Medicine, Faculty of Medicine, Universidad Complutense de Madrid, Plaza de Ramón y Cajal, s/n, 28040 Madrid, Spain; Emergency Department, Hospital General Universitario Gregorio Marañón, Calle del Dr. Esquerdo, 46, 28007 Madrid, Spain; Microbiology Service, Hospital Universitario Rio Hortega, Calle Dulzaina, 2, 47012 Valladolid, Spain; Viral Infection and Immunity Unit, Centro Nacional de Microbiología, Instituto de Salud Carlos III, Ctra. de Pozuelo, 28, 28222 Majadahonda, Spain; Hospital Universitario Infanta Leonor, Av. Gran Vía del Este, 80, 28031 Madrid, Spain; Servicio de Microbiología Clínica, Hospital Universitario Príncipe de Asturias, Carr. de Alcalá, s/n, 28805, Madrid, Spain; Intensive Care Unit, Hospital Universitario Rio Hortega, Calle Dulzaina, 2, 47012 Valladolid, Spain; Emergency Department, Hospital Universitario Rio Hortega, Calle Dulzaina, 2, 47012 Valladolid, Spain; Intensive Care Unit, Hospital Clínico Universitario de Valladolid. Av. Ramón y Cajal, 3, 47003 Valladolid, Spain; Department of Anesthesiology, Facultad de Medicina de Valladolid, Av. Ramón y Cajal, 7, 47005 Valladolid, Spain; Anesthesiology and Reanimation Service, Hospital Universitario Rio Hortega, Calle Dulzaina, 2, 47012 Valladolid, Spain; Intensive Care Unit. Hospital General Universitario Gregorio Marañón. Calle del Dr. Esquerdo, 46, 28007 Madrid, Spain; Microbiology Service, Hospital Clinico Universitario de Valladolid, Av. Ramón y Cajal, 3, 47003 Valladolid, Spain; Pulmonology Service, Hospital Universitario y Politécnico de La Fe, Avinguda de Fernando Abril Martorell, 106, 46026, Valencia Spain; Clinical Microbiology Department. Complejo Asistencial Universitario de León. Calle Altos de nava, s/n, 24001 León, Spain; Microbiology Service, Hospital Universitario de Burgos, Av. Islas Baleares, 3, 09006 Burgos, Spain; Intensive Care Unit. Complejo Asistencial Universitario de León. Calle Altos de nava, s/n, 24001 León, Spain; Pneumology Service, Hospital Universitario Río Hortega / Biomedical Engineering Group, Universidad de Valladolid, Calle Dulzaina, 2, 47012 Valladolid, Spain; Intensive Care Unit. Hospital Universitario de Burgos, Av. Islas Baleares, 3, 09006 Burgos, Spain; Clinical Analysis Service. Hospital Clínico Universitario de Valladolid, Av. Ramón y Cajal, 3, 47003 Valladolid, Spain; Department of Microbiology and Immunology, Faculty of Medicine, Canadian Center for Vaccinology CCfV, Dalhousie University, Halifax, Nova Scotia, B3H 4R2, Canada; Laboratory of Immunity, Shantou University Medical College, 22 Xinling Rd, Jinping, Shantou, Guangdong, China; Department of Cardiovascular Surgery, Hospital Clínico Universitario de Valladolid. Av. Ramón y Cajal, 3, 47003 Valladolid, Spain; Infectious diseases clinic, Internal Medicine Department, Hospital Universitario Río Hortega, Valladolid, Calle Dulzaina, 2, 47012 Valladolid, Spain; Department of Pulmonology, Hospital Clinic de Barcelona, Universidad de Barcelona, Institut D investigacions August Pi I Sunyer (IDIBAPS), Carrer del Rosselló, 149, 08036 Barcelona, Spain; Intensive Care Department, Vall d’Hebron Hospital Universitari. SODIR Research Group, Vall d’Hebron Institut de Recerca, Passeig de la Vall d’Hebron, 119, 08035 Barcelona, Spain; Respiratory Department, Institut Ricerca Biomedica de Lleida, Av. Alcalde Rovira Roure, 80, 25198 Lleida, Spain

**Keywords:** SARS-CoV-2, cytokine, sepsis, COVID-19, plasma, RNAemia, viral RNA load, ICU

## Abstract

**Background:** COVID-19 can course with respiratory and extrapulmonary disease. SARS-CoV-2 RNA is detected in respiratory samples but also in blood, stool and urine. Severe COVID-19 is characterised by a dysregulated host response to this virus. We studied whether viral RNAemia or viral RNA load in plasma are associated to severe COVID-19 and also to this dysregulated response.

**Methods:** 250 patients with COVID-19 were recruited (50 outpatients, 100 hospitalised ward patients, and 100 critically ill). Viral RNA detection and quantification in plasma was performed using droplet digital PCR, targeting the N1 and N2 regions of the SARS-CoV-2 nucleoprotein gene. The association between SARS-CoV-2 RNAemia and viral RNA load in plasma with severity was evaluated by multivariate logistic regression. Correlations between viral RNA load and biomarkers evidencing dysregulation of host response were evaluated by calculating the Spearman correlation coefficients.

**Results:** the frequency of viral RNAemia was higher in the critically ill patients (78%) compared to ward patients (27%) and outpatients (2%) (*p*<0.001). Critical patients had higher viral RNA loads in plasma than non-critically ill patients, with non survivors showing the highest values. When outpatients and ward patients were compared, viral RNAemia did not show significant associations in the multivariate analysis. In contrast, when ward patients were compared with ICU patients, both viral RNAemia and viral RNA load in plasma were associated with critical illness (OR [CI 95%], *p*): RNAemia (3.92 [1.183 - 12.968], 0.025), viral RNA load (N1) (1.962 [1.244 - 3.096], 0.004); viral RNA load (N2) (2.229 [1.382 - 3.595], 0.001). Viral RNA load in plasma correlated with higher levels of chemokines (CXCL10, CCL2), biomarkers indicative of a systemic inflammatory response (IL-6, CRP, Ferritin), activation of NK cells (IL-15), endothelial dysfunction (VCAM-1, angiopoietin-2, ICAM-1), coagulation activation (D-Dimer and INR), tissue damage (LDH, GPT), neutrophil response (neutrophils counts, myeloperoxidase, GM-CSF) and immunodepression (PD-L1, IL-10, lymphopenia and monocytopenia).

**Conclusions:** SARS-CoV-2 RNAemia and viral RNA load in plasma are associated to critical illness in COVID-19. Viral RNA load in plasma correlates with key signatures of dysregulated host responses, suggesting a major role of uncontrolled viral replication in the pathogenesis of this disease.

## Background

With well over 43 million cases and 1.56212 deaths globally, Coronavirus disease 2019 (COVID-19) has become the top economic and health priority worldwide [1]. Among hospitalized patients, around 10–20% are admitted to the intensive care unit (ICU), 3–10% require intubation and 2–5% die [2]. SARS-CoV-2 RNA is commonly detected in nasopharyngeal swabs; however viral RNA can be found in sputum, lung samples, peripheral blood, serum, stool samples, and to a limited extent urine [3] [4] [5] [6]. While the lungs are most often affected, severe COVID-19 also induce inflammatory cell infiltration, haemorrhage, and degeneration or necrosis in extra-pulmonary organs (spleen, lymph nodes, kidney, liver, central nervous system) [7] [8]. Patients with severe COVID-19 show signatures of dysregulated response to infection, with immunological alterations involving moderate elevation of some cytokines and chemokines such as IL-6, IL-10 or CXCL10, deep lymphopenia with neutrophilia, systemic inflammation (elevation of C Reactive Protein, ferritin), endothelial dysfunction, coagulation hyper-activation (D-dimers) and tissue damage (LDH) [9] [10] [11] [12] [13] [14].

Our hypothesis is that systemic distribution of the virus or viral components could be associated to the severity of COVID-19, and, in turn to a number of parameters indicating the presence of a dysregulated response to the infection.

While the SARS-CoV-2 virus has been reported to be difficult to culture from blood [4], PCR based methods are able to detect and quantify the presence of genomic material of the virus in serum or plasma, representing an useful approach to evaluate the impact of the extrapulmonary dissemination of viral material on disease severity and also on the host response to the infection [5] [15]. An excellent approach for achieving absolute quantification of viral RNA load is droplet digital PCR (ddPCR). ddPCR is a next-generation PCR method, which offers absolute quantification with no need of standard curve and greater precision and reproducibility than currently available qRT-PCR methods, as revised elsewhere [16].

We employed here ddPCR to detect and quantify viral RNA in plasma from COVID-19 patients discharged from the emergency room with mild severity, patients admitted to the ward with moderate severity, and critically ill patients. Our objectives in this study were: 1) to evaluate if there is an association between SARS-CoV-2 RNAemia and viral RNA load with moderate disease; 2) to evaluate if there is an association between SARS-CoV-2 RNAemia and viral RNA load with critical illness; 3) to evaluate the correlations between SARS-CoV-2 RNA load in plasma and parameters of dysregulated host responses against SARS-CoV-2.

## Methods

### Study design

250 adult patients with a positive nasopharyngeal swab polymerase chain reaction (PCR) test for SARS-CoV-2 performed at participating hospitals were recruited during the first pandemic wave in Spain from March 16^th^ to the 15^th^ of April 2020. The patients recruited were of three different categories. The first corresponded to patients examined at an emergency room and discharged within the first 24 hours (outpatients group, n=50). The second group were patients hospitalized to pneumology, infectious diseases or internal medicine wards (wards group, n=100). Patients who required critical care or died during hospitalization were excluded from this group, in order to have a group of clear moderate severity. The third group corresponded to patients admitted to the ICU (n=100). Patient’s recruited by participating hospital are detailed in the additional file 1. 20 healthy blood donors were included as controls. These controls were recruited during the pandemics, in parallel to the SARS-CoV-2 infected patients, and were negative for SARS-CoV-2 IgG. This study was registered at Clinicaltrials.gov with the identification NCT04457505.

### Blood samples

Plasma from blood collected in EDTA tubes samples was obtained from the three groups of patients in the first 24 hours following admission to the emergency room, to the ward, or to the ICU, at a median collection day since disease onset of 7, 8 and 10 respectively, and also. from 20 blood donors (10 men and 10 women).

### Biomarker profiling

a panel of biomarkers was profiled by using the Ella-SimplePlex™ immunoassay (San Jose, California, U.S.A), informing of the following biological functions potentially altered in severe COVID-19, based in the available evidence on COVID-19 physiopathology [13] [17] and also in our previous experience on emerging infections and sepsis [18] [19] [20] [21]: neutrophil degranulation: Lipocalin-2/NGAL, myeloperoxidase; endothelial dysfunction: ICAM-1, VCAM-1/CD106, Angiopoietin 2; T cell survival and function: IL-7, Granzyme B; immunosuppression: IL-1ra, B7-H1/PD-L1, IL-10; chemotaxis: CXCL10/IP10, CCL2; Th1 response: Interleukin 1 beta, IFN-γ, IL-12p70, IL-15, TNF-α, IL-2; Th2 response: IL-4, IL-10; Th17 response: IL-6, IL-17A; granulocyte mobilization / activation: G-CSF, GM-CSF; coagulation activation: D-Dimer; acute phase reactants: Ferritin (C Reactive protein and LDH were profiled in each participant hospital by their central laboratories).

### Detection and quantification of SARS-CoV-2 RNA in plasma

RNA was extracted from 100 µl of plasma using an automated system, eMAG® from bioMérieux® (Marcy l’Etoile, France). Detection and quantification of SARS-CoV-2 RNA was performed in five µl of the eluted solution using the Bio-Rad SARS-CoV-2 ddPCR kit according to manufacturer’s specifications on a QX-200 droplet digital PCR platform from the same provider. This PCR targets the N1 and N2 regions of the viral nucleoprotein gene and also the human ribonuclease (RNase) P gene using the primers and probes sets detailed in the CDC 2019-Novel Coronavirus (2019-nCoV) Real-Time RT-PCR Diagnostic Panel [22]. Samples were considered positive for SARS-CoV-2 when N1 and/or N2 presented values ≥ 0.1 copies/µL in a given reaction. RNase P gene was considered positive when it presented values ≥ 0.2 copies/µL, following manufacturer`s indications. The test was only considered valid when RNase P gene was positive. Final results were given in copies of cDNA / mL of plasma. IgG specific for the Nucleocapsid Protein of SARS–CoV-2 was detected in 150 µl of plasma using the Abbott Architect SARS-CoV-2 IgG Assay (Illinois, U.S.A). Viral RNA and SARS-CoV-2 IgG were profiled in the same plasma sample.

### Statistical analysis

For the demographic and clinical characteristics of the patients, the differences between groups were assessed using the Chi-square test / Fisher’s Exact Test where appropriated for categorical variables. Differences for continuous variables were assessed by using the Kruskal-Wallis test with post hoc tests adjusting for multiple comparisons. Multivariate logistic regression analysis was employed to evaluate the association between viral RNAemia and viral RNA load in plasma with severity, in the comparisons [outpatients vs ward patients] and [ward patients vs ICU patients]. Variables showing significant differences between groups in each comparison in the Kruskal-Wallis test were further introduced in the multivariate analysis as adjusting variables. The list of variables considered as potential adjusting variables were [Age (years)], [Sex (male)], [Alcoholism], [Smoker], [Drug abuse], [Cardiac disease], [Chronic vascular disease], [COPD], [Asthma], [Obesity], [Hypertension], [Dyslipidemia], [Chronic renal disease], [Chronic hepatic disease], [Neurological disease], [HIV], [Autoimmune disease], [Chronic inflammatory bowel disease], [Type 1 diabetes], [Type 2 diabetes], [Cancer], [Invasive mechanical ventilation], [Non-invasive mechanical ventilation], [SARS-CoV-2 IgG], [Temperature (°C)], [Systolic Pressure (mmHg)], [Oxygen saturation (%)], [Bilateral pulmonary infiltrate], [Glucose (mg/dl)], [Creatinine (mg/dl)], [Na (mEq/L)], [K (mEq/L)], [Platelets (cell × 10^3^ / µl)], [INR], [D Dimer (pg/ml)], [LDH (UI/L)], [GPT (UI/L)], [Ferritin (pg/ml)], [CRP (mg/dl)], [Haematocrit (%)], [Lymphocytes (cells/mm3)], [Neutrophils (cells/mm3)], [Monocytes (cells/mm3)]. Multivariate logistic regression analysis was performed using the “Enter” method, but also the backward stepwise selection method (Likelihood Ratio) was employed in each case to confirm the association between viral RNAemia and viral RNA load in plasma with disease severity (pin < 0.05, pout < 0.10), not forcing entry of these variables in the model. Correlation analysis were performed using the Spearman test applying the Bonferroni correction of the *p* value. Variables evaluated for correlation with viral RNA load were: [Temperature (°C)], [Systolic Pressure (mmHg)], [Oxygen saturation (%)], [Lymphocytes (cells/mm3)], [Neutrophils (cells/mm3)], [Monocytes (cells/mm3)], [Creatinine (mg/dl)], [LDH (UI/L)], [GPT (UI/L)], [Platelets (cell × 10^3^ / µl)], [INR], [CRP (mg/dl)], and all the biomarkers analyzed by Ella-SimplePlex. Statistical analysis was performed with IBM SPSS® version 20 (IBM, Armonk, New York, USA).

## Results

### Clinical characteristics of the patients (Table 1)

Patients requiring hospitalization (either general ward or ICU) were older than those patients discharged to their home from the ER. Critically ill patients were more frequently male than those in the other groups. Comorbidities of obesity, hypertension, dyslipidemia and type 2 diabetes were more commonly found in patients requiring hospitalization, with no significant differences found in the comorbidities profile between critically ill and non-critically ill hospitalized patients. 14% of the patients in clinical wards required non-invasive mechanical ventilation, while 96% of the patients admitted to the ICU required invasive mechanical ventilation. Critically ill patients had increased glucose levels, along with higher concentration of neutrophils in blood, increased levels of ferritin and C-reactive protein (denoting activation of the systemic inflammatory response). Increased levels of INR and D-dimers (reflecting activation of the coagulation system), as well as LDH and GPT, which levels raise as consequence of tissue and liver damage, were also observed in critically ill patients. Patients admitted to the ICU also showed a lower haematocrit, pronounced lymphopenia and lower monocyte counts at admission. ICU patients stayed longer in the hospital than ward patients, with 49 % having a fatal outcome.

**Table 1:**
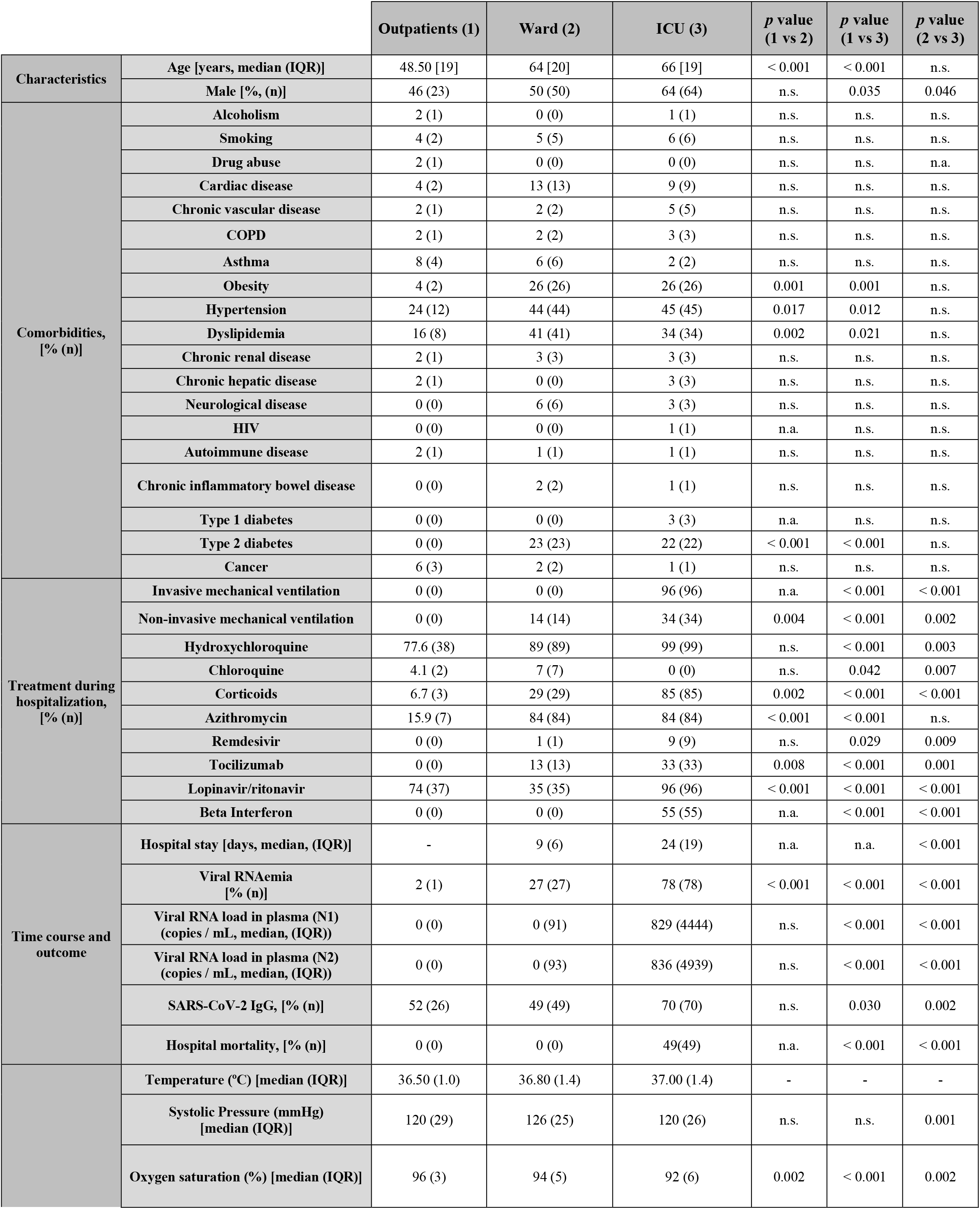
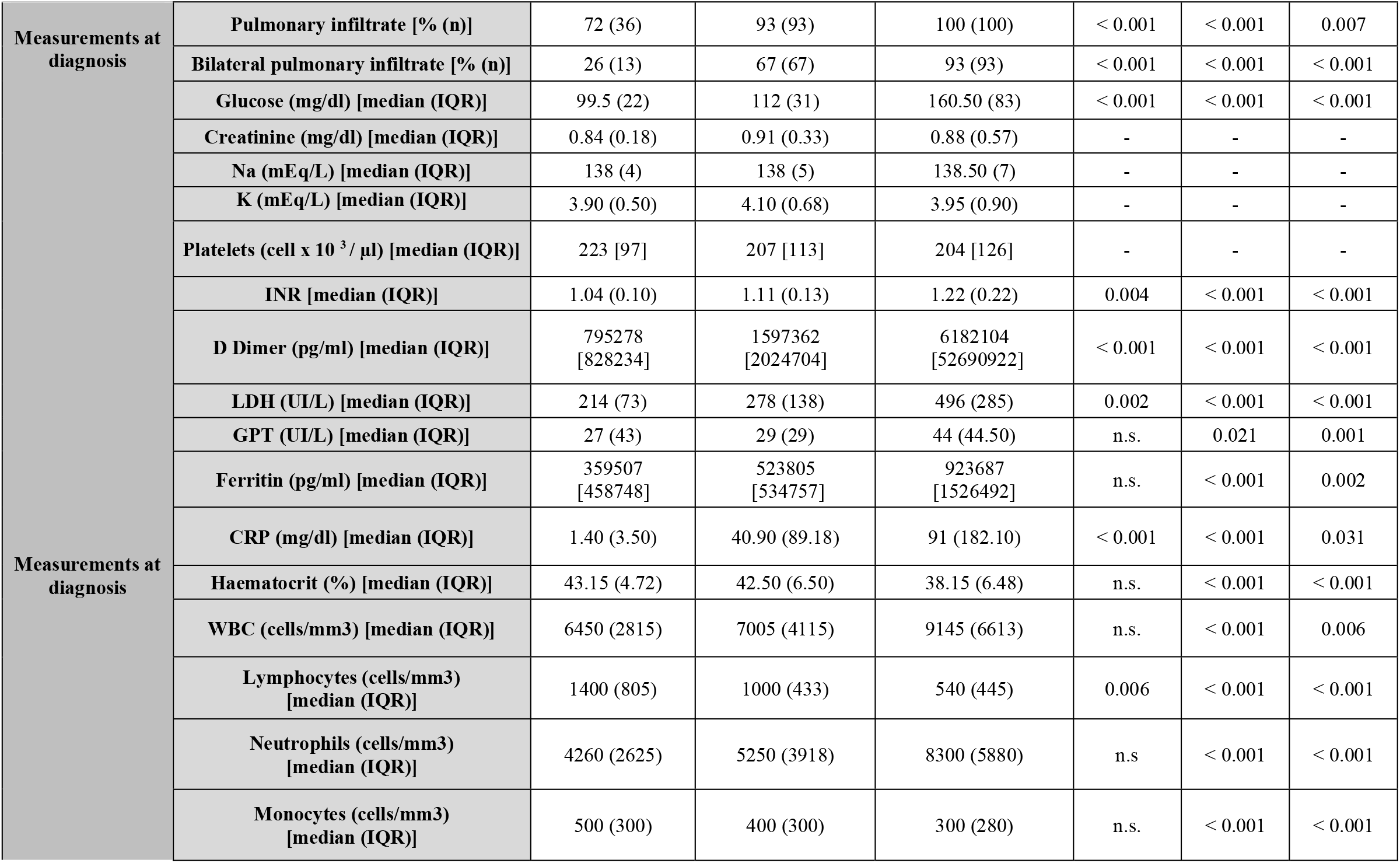
Clinical characteristics of the patients: continuous variables are represented as [median, (interquartile range, IQR)]; categorical variables are represented as [%, (n)].; INR, International Normalized Ratio; n.s., not significant; n.a., not applicable. COPD (Chronic obstructive pulmonary disease), HIV (Human Immunodeficiency Virus), INR (International Normalized Ratio), LDH (Lactic Acid Dehydrogenase), GPT (glutamic-pyruvate transaminase); CRP (C-reactive protein), WBC (white blood cell).

### Viral RNAemia, viral RNA load in plasma and specific SARS-Cov-2 IgG in the three groups of patients

As depicted in table 1, the frequency of the detection of SARS-CoV-2 viral RNA (RNAemia) was significantly higher in the critically ill patients (78%) compared to ward patients (27%) and outpatients (2%) (*p*<0.001). Similarly, the group of critically patients showed higher viral RNA loads in plasma than either ward or outpatients (*p*<0.001) (Table 1 and Figure 1). Non survivors showed the highest concentrations of viral RNA in plasma: viral RNA load (N1 region) in ICU non survivors: 1587 copies / ml [10248]; viral RNA load (N1 region) in ICU survivors 574 copies / mL [1872] (results expressed as median [interquartile rank]); viral RNA load (N2 region) in ICU non survivors: 2798 copies / ml [12012]; viral RNA load (N2 region) in ICU survivors 523 copies / mL [1478]. Patients admitted to the wards showed a significant higher frequency of viral RNAemia than outpatients, but viral RNA loads were not significantly different with the latter group (Table 1 and Figure 1). Critically ill patients also had a higher frequency of specific SARS-CoV-2 IgG responses than the other groups (70% in ICU compared to 52% and 49% in the outpatients and ward groups, *p* < 0.05, table 1). No significant differences were found between the group of outpatients and those admitted to the ward. The prevalence of viral RNAemia did not differ between those patients testing positive and those testing negative for SARS-CoV-2 IgG (43.8 % and 40.4 % respectively, *p* =0.586), who in addition showed no differences in viral RNA load (data not shown). Patients with viral RNAemia showed no differences in the days since onset of symptoms compared to those with no viral RNAemia (8.0 days [6.0]; 8.0 days [7.2], *p* = 0.965). In contrast, samples from patients with SARS-CoV-2 IgG were collected later since disease onset that those without SARS-CoV-2 IgG (10.0 days [7]; 7.0 days [6.0], *p* = 0.003)

**Figure 1:**
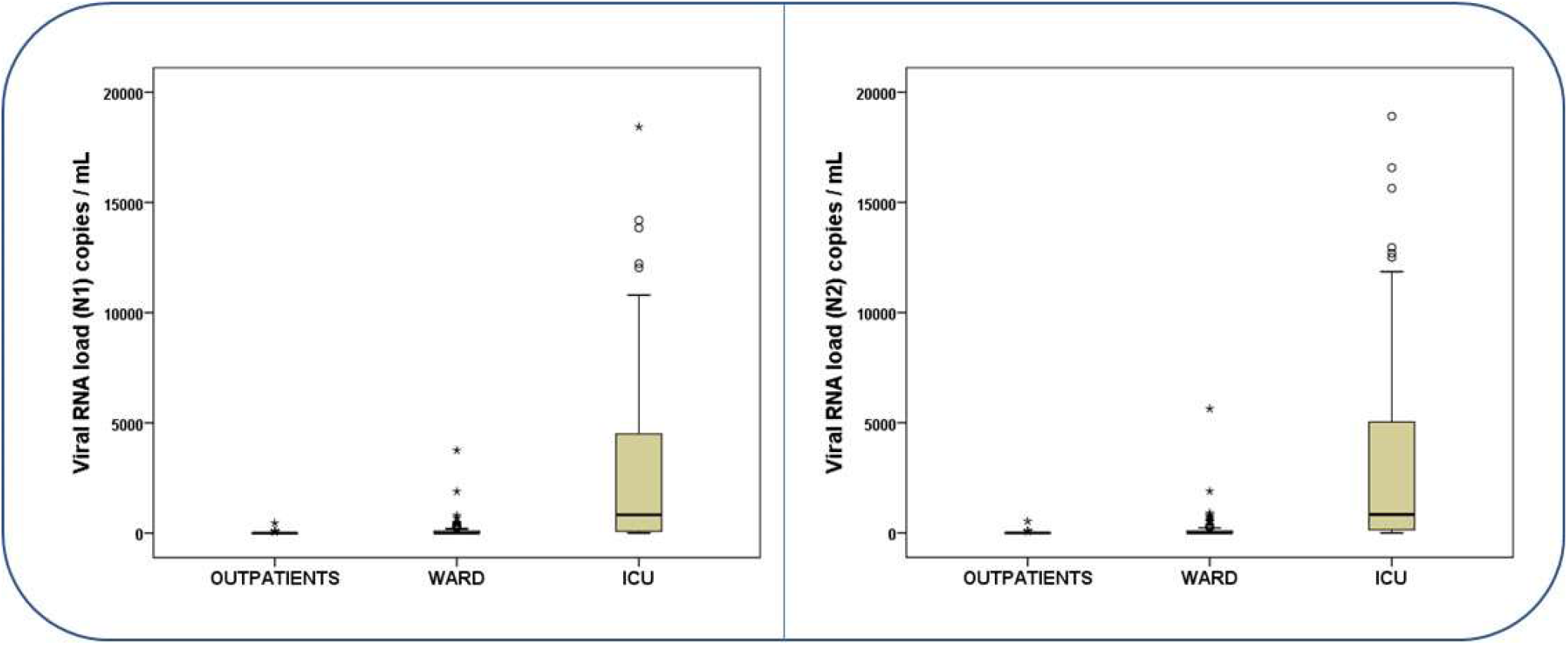
Viral RNA load in plasma, targeting the N1 region (left) and the N2 region (right), in the three groups of patients. Results are provided as copies of cDNA per mL of plasma.

### Multivariate analysis to evaluate the association between viral RNAemia and viral RNA load in plasma with moderate disease and critical illness

While the proportion of patients with viral RNAemia was higher in the wards group compared to the outpatients’ group (table 1), the multivariate analysis did not show a significant association between the presence of viral RNAemia and being hospitalised at the ward, with none of the both methods employed (Additional files 2 and 3). In contrast, when the ward group was compared with critically ill patients, a significant direct association was found between viral RNAemia and viral RNA load in critically ill patients, using the “Enter” method (Table 2) but also the backward stepwise selection method (additional file 4).

**Table 2:** Multivariate logistic regression analysis comparing wards patients against critically ill patients (Enter method). The association between viral RNAemia or viral RNA load targeting the N1 region, or viral RNA load targeting the N2 region with critical illness was evaluated adjusting by major confounding factors.

|  | OR [CI95%] | *p* | OR [CI95%] | *p* | OR [CI95%] | *p* |
| --- | --- | --- | --- | --- | --- | --- |
| Sex (male) | 2.055 [0.656 - 6.432] | 0.216 | 1.647 [0.508 - 5.337] | 0.406 | 1.608 [0.500 - 5.166] | 0.425 |
| Systolic Pressure (mmHg) | 0.985 [0.960 - 1.011] | 0.253 | 0.984 [0.958 - 1.011] | 0.247 | 0.984 [0.957 - 1.012] | 0.261 |
| O2 Saturation (%) | 1.069 [0.986 - 1.160] | 0.107 | 1.076 [0.992 - 1.167] | 0.079 | 1.082 [0.993 - 1.180] | 0.071 |
| Bilateral pulmonary infiltrate | 2.343 [0.469 - 11.707] | 0.300 | 3.088 [0.566 - 16.852] | 0.193 | 3.064 [0.539 - 17.430] | 0.207 |
| Glucose (mg/dl) | 1.008 [1.001 - 1.015] | 0.028 | 1.007 [1.000 - 1.015] | 0.061 | 1.007 [0.999 - 1.014] | 0.070 |
| INR | 1.123 [0.433 - 2.915] | 0.812 | 1.245 [0.469 - 3.305] | 0.661 | 1.377 [0.519 - 3.657] | 0.521 |
| D-dimer (pg/mL) | 1.000 [1.000 - 1.000] | 0.294 | 1.000 [1.000 - 1.000] | 0.190 | 1.000 [1.000 - 1.000] | 0.420 |
| LDH (UI/L) | 1.009 [1.004 - 1.014] | 0.000 | 1.008 [1.003 - 1.013] | 0.002 | 1.009 [1.004 - 1.014] | 0.001 |
| GPT (UI/L) | 1.005 [0.992 - 1.019] | 0.443 | 1.005 [0.991 - 1.019] | 0.516 | 1.006 [0.992 - 1.021] | 0.402 |
| Ferritin (pg/mL) | 1.000 [1.000 - 1.000] | 0.705 | 1.000 [1.000 - 1.000] | 0.856 | 1.000 [1.000 - 1.000] | 0.835 |
| CRP (mg/dl) | 0.998 [0.992 - 1.003] | 0.420 | 0.999 [0.993 - 1.005] | 0.749 | 0.998 [0.993 - 1.004] | 0.547 |
| Haematocrit (%) | 0.770 [0.676 - 0.877] | 0.000 | 0.762 [0.665 - 0.872] | 0.000 | 0.737 [0.633 - 0.858] | 0.000 |
| SARS-CoV-2 IgG (Yes) | 1.967 [0.627 - 6.170] | 0.246 | 1.835 [0.562 - 5.987] | 0.315 | 1.918 [0.577 - 6.377] | 0.288 |
| Lymphocytes (cells/mm3) | 0.998 [0.996 - 1.000] | 0.011 | 0.998 [0.996 - 1.000] | 0.012 | 0.998 [0.996 - 0.999] | 0.010 |
| Monocytes (cells/mm3) | 0.998 [0.995 - 1.001] | 0.180 | 0.998 [0.995 - 1.001] | 0.212 | 0.999 [0.996 - 1.002] | 0.463 |
| Neutrophils (cells/mm3) | 1.000 [1.000 - 1.000] | 0.262 | 1.000 [1.000 - 1.000] | 0.326 | 1.000 [1.000 - 1.000] | 0.239 |
| **Viral RNAemia (Yes)** | **3.916 [1.183 - 12.968]** | **0.025** | - | - | - | - |
| **Viral RNA load (N1)**<br>**in plasma, log (copies/mL)** | - | - | **1.962 [1.244 - 3.096]** | **0.004** | - | - |
| **Viral RNA load (N2)**<br>**in plasma, log (copies/mL)** | - | - | - | - | **2.229 [1.382 - 3.595]** | **0.001** |

### Correlations between viral RNA load in plasma and biological responses to SARS-CoV-2 infection

viral RNA load in plasma (targeting either the N1 and the N2 regions) showed the strongest direct correlations with plasma levels of CXCL10, LDH, IL-10, IL-6, IL-15, myeloperoxidase and CCL-2 (MCP-1) and inverse correlations with lymphocytes, monocytes and O2 saturation (Figure 2). These were the parameters whose levels varied the most in critically patients compared with ward and outpatient groups (Figures 3 and 4 and additional file 5). CXCL10 was the most accurate identifier of viral RNAemia in plasma (area under the curve (AUC), [CI95%], p) = 0.85 [0.80 – 0.89), <0.001), and IL-15 was the cytokine which most accurately differentiated clinical ward patients from ICU patients (AUC: 0.82 [0.76 – 0.88], <0.001). Plasma viral RNA load also showed significant direct correlations with levels of VCAM-1, PDL-1, GM-CSF, G-CSF, neutrophil counts, IL-1ra, CRP, INR, D-dimer, TFNα, Angiopoietin-2, GPT, ICAM-1, IL-7 and Ferritin (Figure 2), with most of these mediators showing the highest variations in the critically ill patients (Figures 3 and 4 and additional file 5).

**Figure 2.**
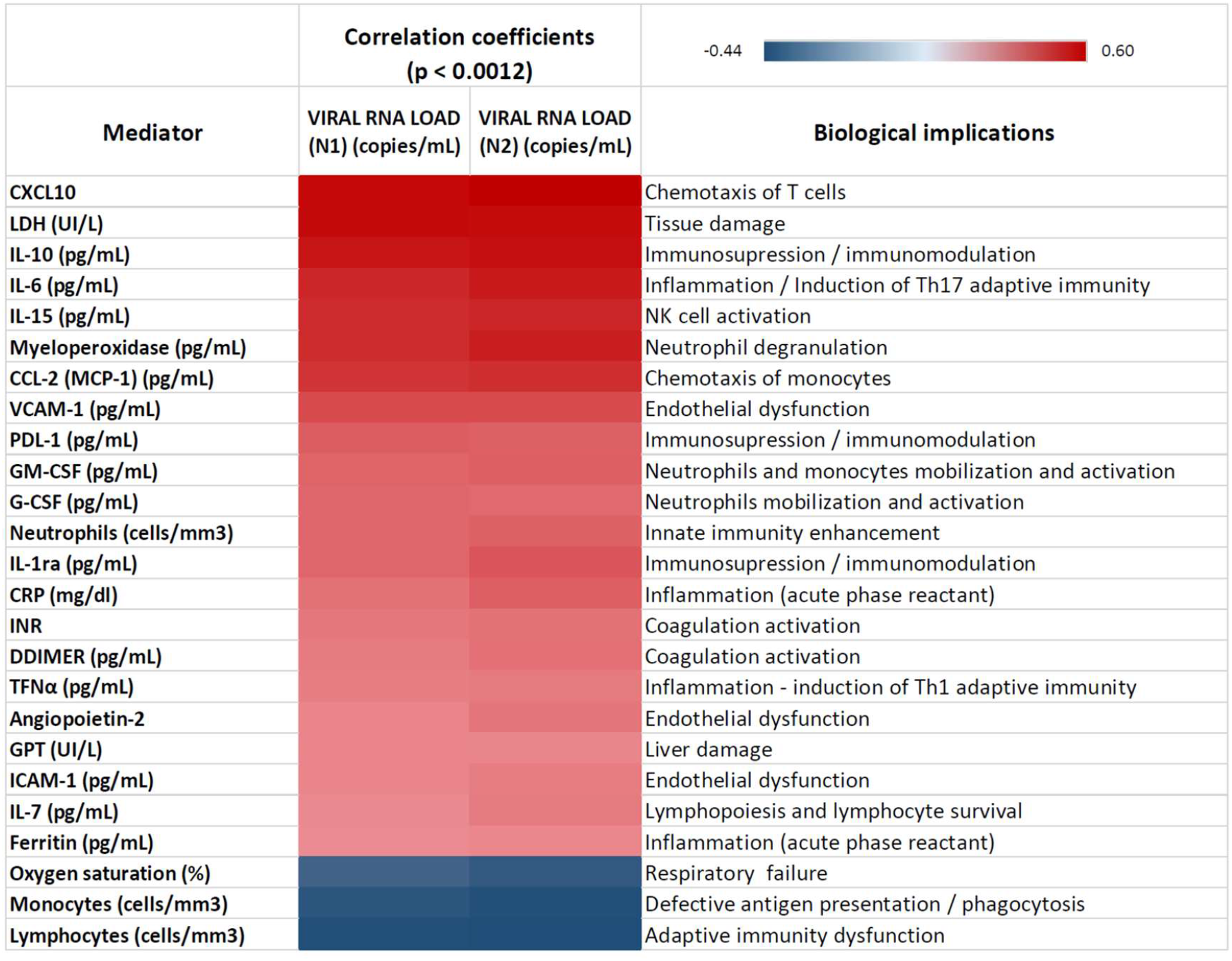
Heat map representing the Spearman correlation coefficients between viral RNA load in plasma targeting the N1 and the N2 regions and representative indicators of host dysregulated response.

**Figure 3.**
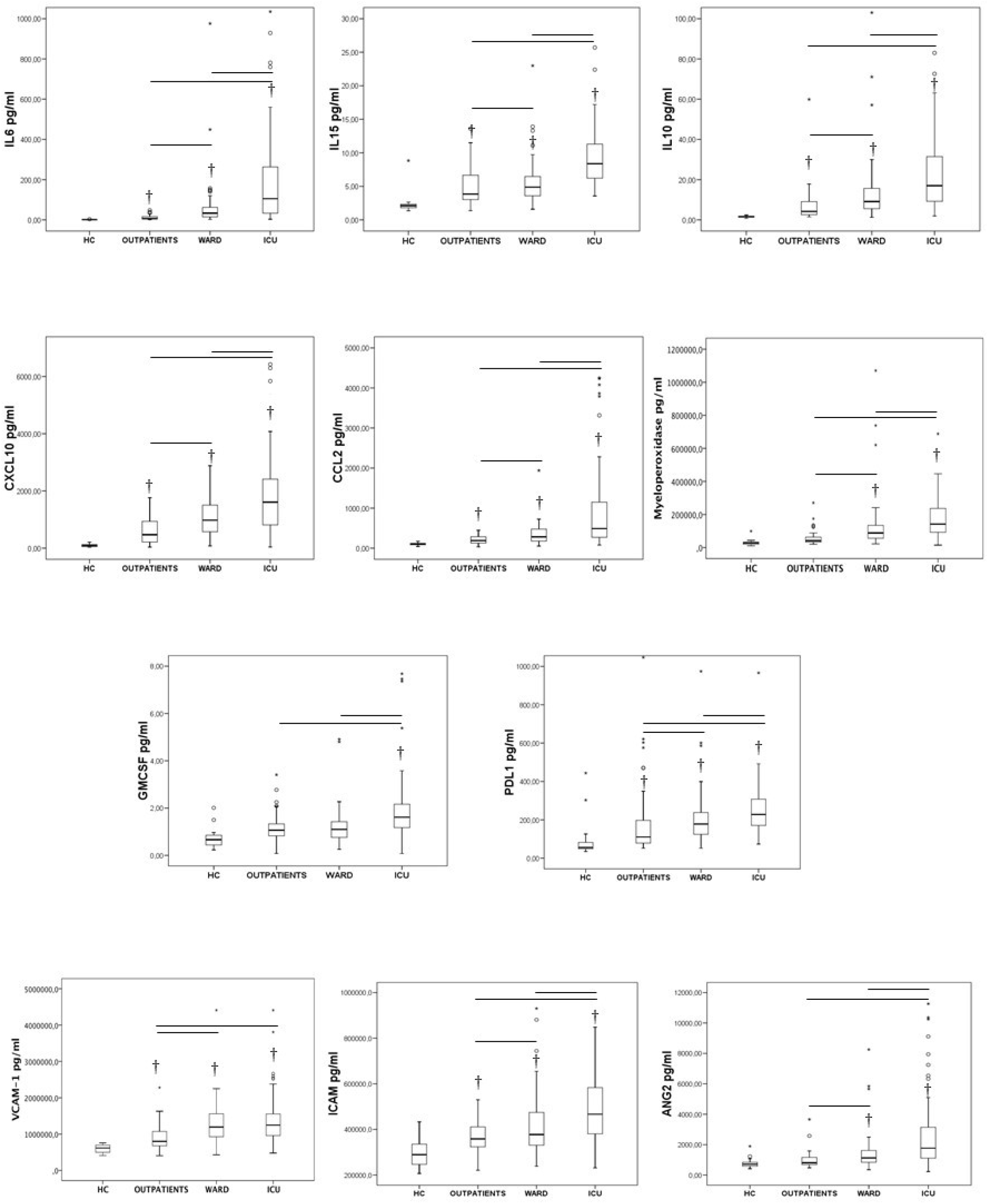
Levels of laboratory parameters indicating host dysregulated response across groups. † indicates significant difference with the healthy control and the bars significant differences between the other groups.

**Figure 4:**
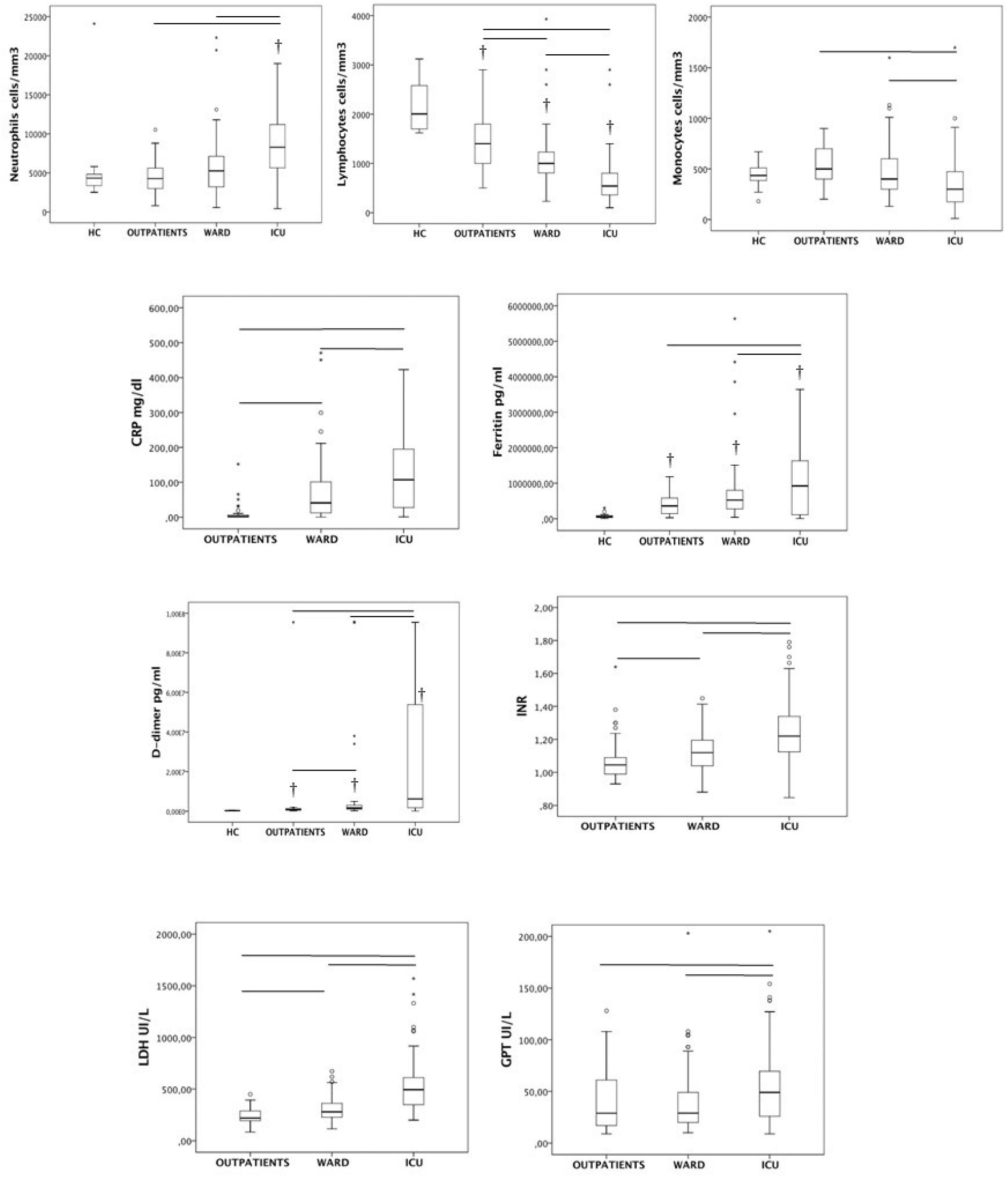
Levels of laboratory parameters indicating host dysregulated response across groups. † indicates significant difference with the healthy control and the bars significant differences between the other groups.

## Discussion

Our study demonstrates that the presence of SARS-CoV-2-RNA in plasma is associated to critical illness in COVID-19 patients, with the strength of association being the highest in those patients with the highest viral RNA loads. This association was independent of other factors also related to disease severity. Moreover, those critically ill patients who died presented with higher viral RNA loads in plasma than those who survived. SARS-CoV-2 viral RNA was detected in the plasma of the vast majority of those COVID-19 patients admitted to the ICU (78%). As far as we know, our study is the largest one to date using ddPCR to quantify SARS-CoV-2 RNA load in plasma from COVID-19 patients, and the only one with a multicentric design. Our results are in consonance with those from Veyer *et al*, who, in a pilot study using this technology, found higher viral RNA loads and a prevalence of RNA viremia of 88% in twenty six COVID-19 patients who were critically ill [16]. The results are also in agreement with those of Hagman et al, who, using standard RT-PCR technology, found that the presence of SARS-CoV-2 RNA in serum at hospital admission was associated with a seven-fold increased risk of critical disease and an eight-fold increased risk of death in a cohort of 167 patients hospitalised for COVID-19 [23].

Although our study did not determine if the presence of viral RNA in plasma reflects the presence of live virus in peripheral blood, the association found between the presence and concentration of viral RNA in plasma and critical illness suggests that viral replication is more robust in severe COVID-19, and/or that critically ill patients with this disease are not able to control viral replication. This notion is further supported by the correlations found in our study between viral RNA load in plasma and hypercytokinemia involving CXCL10, IL-10, CCL2, IL-6 and IL-15, where the levels of these cytokines were the highest in patients with critical illness. The correlation between viral RNA load and higher levels of cytokines has also been described in the severe infections caused by H5N1 and pandemic H1N1 influenza strains [24] [25]. Active viral replication stimulates the secretion of cytokines by the recognition of viral RNA by endosomal receptors such as toll like receptor 7 (TLR7) in human plasmacytoid dendritic cells and B cells, or TLR8 in myeloid cells [26]. While the elevation of CXCL10, IL-10, CCL2, IL-6 has been extensively documented in severe COVID-19 [9] [10], our work demonstrates a clear correlation between these cytokines and plasma viral load. Furthermore, we report for the first time a major role of IL-15 in severe COVID-19. High levels of IL-15 in critically ill patients with high SARS-CoV-2 RNA load in plasma could be an attempt to stimulate Natural Killer cells to fight the virus [27]. We previously demonstrated that high levels of IL-15, along with IL-6, constituted a signature of critical illness in H1N1 pandemic influenza infection [20].

Viral RNA load correlated with higher levels of myeloperoxidase in plasma, which were the highest in those patients admitted to the ICU. This is a marker of neutrophil degranulation and a potent tissue damage factor which has been proposed to play a role in the pathogenesis of ARDS secondary to influenza, by mediating claudin alteration on endothelial tight junctions, eventually leading to protein leakage and viral spread [28]. In this regard, the correlation found between viral RNA load in plasma and higher levels of LDH and GPT could suggest a direct or indirect role of viral replication in mediating tissue destruction in COVID-19.

Interesting, but less robust, direct correlations were found between viral RNA load in plasma with GM-CSF and neutrophil counts in blood, further reinforcing the role of neutrophil mediated responses in the pathogenesis of severe COVID-19. The direct correlation with soluble PDL-1 is also relevant, since this the ligand of the inhibitory co-receptor PD1 on T cells, which activation induces anergy of T lymphocytes [29]. This finding reinforces the potential role of immune checkpoint inhibitors in severe COVID-19 [30]. In turn, the association found between viral RNA load in plasma and three mediators of endothelial dysfunction (VCAM-1, angiopoietin 2 and ICAM-1), and with coagulation activation markers (D-dimers and INR prolongation) suggests a potential virally linked mechanism in the pathogenesis of endothelitis and thrombosis in COVID-19 disease [7]. Finally, the correlation with the acute phase reactants CRP and ferritin suggests a connection between shedding of genomic material of the virus to the blood and the induction of a systemic inflammatory response which is observed in those patients needing critical care.

The strongest inverse correlations found in our study was between SARS-CoV-2 RNA load in plasma and lymphocyte and monocyte counts in peripheral blood, for which critically ill patients showed the lowest values. Active viral replication could be a precipitating event in the pathogenesis of lymphopenia and monocytopenia in severe COVID-19 patients [11] [31], by mediating direct cytopathic actions or stimulating the migration of these cells to the extravascular space to reach the infected tissues [32].

A limitation of our work is its observational nature, which precludes to infer causality. Nonetheless, the observed associations could serve as hypothesis generators, leading to the development of animal models to confirm the potential link between SARS-CoV-2 replication and the dysregulated host responses observed in severe COVID-19.

## Conclusion

Presence of SARS-CoV-2 RNA in plasma is associated with critical illness in patients with COVID-19. The strength of this association increases with viral RNA load in plasma, which in turn correlates with key signatures of dysregulated host response in COVID-19 (figure 5). Our findings suggest a major role of uncontrolled viral replication in the pathogenesis of this disease. Assessment of viral RNAemia and viral RNA load in plasma could be useful to early detect those patients at risk of clinical deterioration, to assess response to treatment and to predict disease outcome.

**Figure 5:**
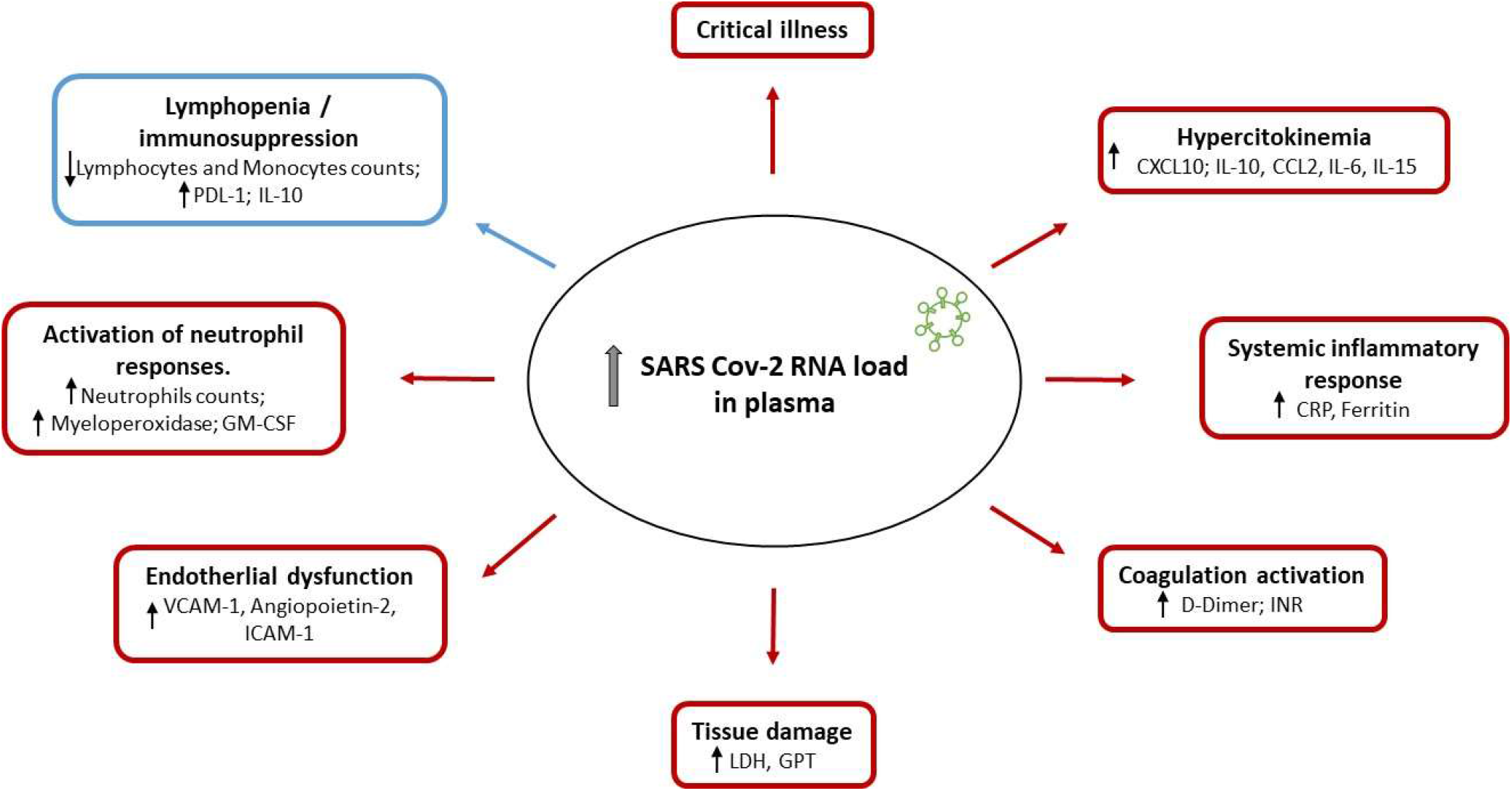
Integrative model depicting the correlations found between the indicators of host dysregulated responses and SARS-CoV-2 RNA load in plasma.

## Data Availability

Data are available upon reasonable request

## List of abbreviations

SARS-CoV-2: Severe acute Respiratory Syndrom-Coronavirus-2
LDH: Lactate dehydrogenase
SF: Granulocyte colony-stimulating factor
TLR: toll like receptor

## Declarations

### Ethics approval and consent to participate

The study was approved by the Committee for Ethical Research of the coordinating institution, “Comite de Etica de la Investigacion con Medicamentos del Area de Salud de Salamanca”, code PI 2020 03 452. Informed consent was obtained orally when clinically possible. In the remaining cases, the informed consent waiver was authorized by the Ethics committee.

### Consent for publication

not applicable

### Availability of data and materials

the datasets generated and/or analysed during the current study are not publicly available since they are still under elaboration for publication by the authors but are available from the corresponding author on reasonable request.

### Authors ‘contribution

JFBM, DJK, JB, RF, FB, AT and RM designed the study. JFBM and DJK wrote the manuscript and interpreted the data. RA coordinated the clinical study and drafted the figures. MGR, DM, PR, FPG, LT, RLI, EB, CA, JMG, JR, RM, MIF, GM, MGE, DC, FDC, JFR, WT, PGJ, GR, IM, EG,IM, SP, SM, PGO, JAC, TRA, CP, JAB, GR, RH, JB, PE, RC, JA, JGB, NM, NBL, LJV, BFC, MAM recruited the patients and /or collected the clinical data. MDG, AO, RO, LMR and JME performed the assays for the detection of SARS-CoV-2 IgG and viremia. APT, CD and AO developed the ddPCR works; CD and NJ profiled the immunological mediators. SR, AMF and MMF developed the statistical analysis and drafted the figures. AAK, ATO, AM and LF performed the literature search. All authors read and approved the final manuscript.

## Ackowledgements

we thank SEIMC-GESIDA Foundation for the scientific sponsoring of this project. We thank also the “Biobanco del Centro de Hemoterapia y Hemodonación de Castilla y León”, which provided the plasma simples used in the healthy control group.

---

**Additional file 1:** recruited patients in each hospital.

| Hospital | Outpatients | Wards | ICU |
| --- | --- | --- | --- |
| Hospital Clínico Universitario de Valladolid | 0 | 0 | 45 |
| Hospital General Universitario Gregorio Marañón (Madrid) | 44 | 4 | 25 |
| Hospital Universitario Infanta Leonor (Madrid) | 0 | 30 | 0 |
| Hospital Universitario y Politécnico de La Fe (Valencia) | 0 | 27 | 0 |
| Hospital Universitario Río Hortega (Valladolid) | 2 | 22 | 23 |
| Hospital Universitario Príncipe de Asturias (Madrid) | 4 | 17 | 1 |
| Hospital Universitario de Burgos | 0 | 0 | 2 |
| Hospital Universitario de León | 0 | 0 | 4 |
| Total | 50 | 100 | 100 |

**Additional file 2.**
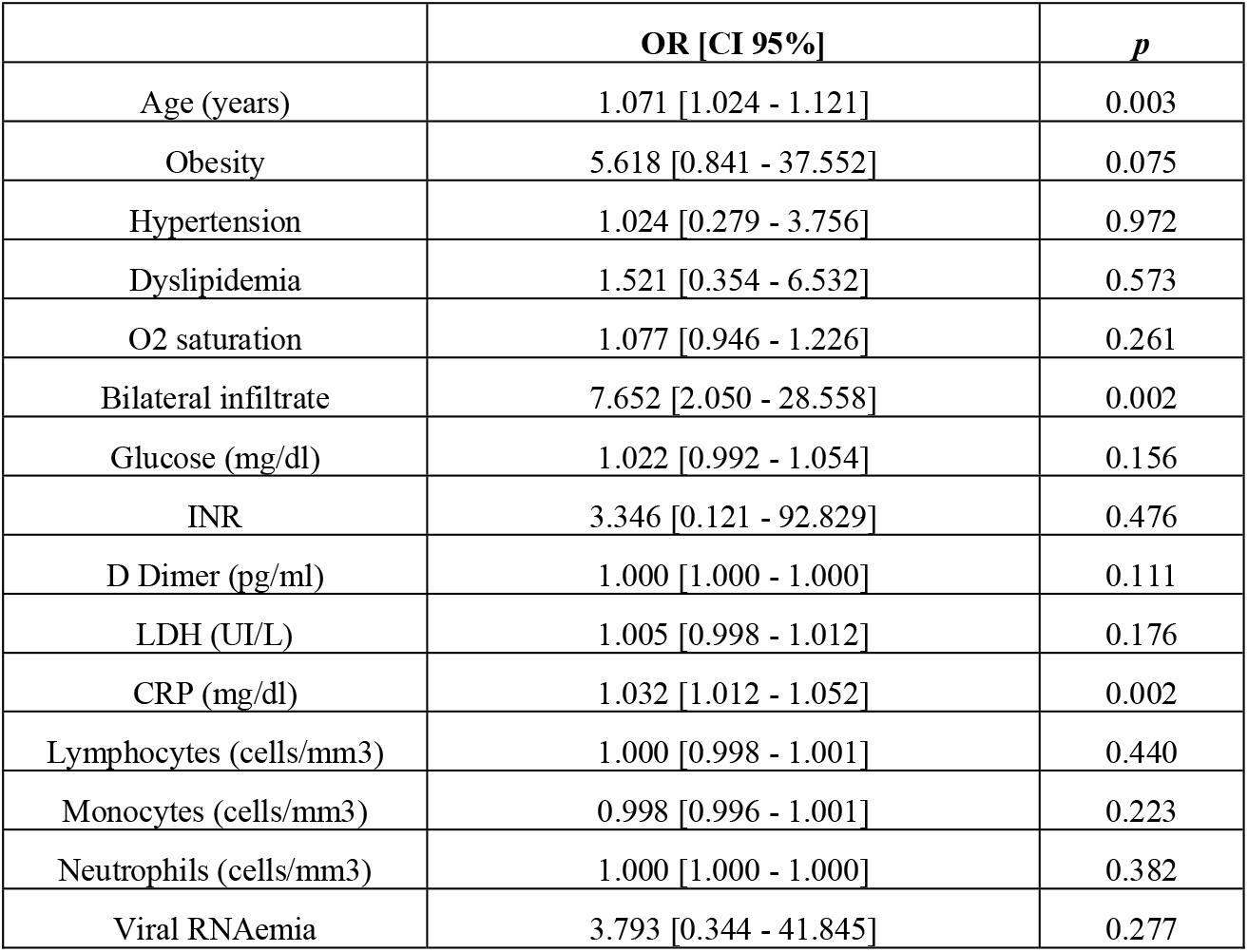
Multivariate logistic regression analysis comparing outpatients against wards patients (Enter method). The association between viral RNAemia with hospitalization at the wards was evaluated adjusting by major confounding factors.

**Additional file 3.** Multivariate logistic regression analysis comparing outpatients against wards patients (backward stepwise selection method / Likelihood Ratio). The association between viral RNAemia with hospitalization at the wards was evaluated adjusting by major confounding factors, but it was not selected in the final model.

|  | OR [CI95%] | *p* |
| --- | --- | --- |
| Age | 1.057 [1.017 - 1.098] | 0.005 |
| Obesity | 6.675 [1.129 - 39.472] | 0.036 |
| Bilateral infiltrate | 6.400 [2.153 - 19.024] | 0.001 |
| Glucose (mg/dl) | 1.027 [0.999 - 1.055] | 0.058 |
| CRP (mg/dl) | 1.032 [1.013 - 1.051] | 0.001 |
| Monocytes (cells/mm3) | 0.997 [0.995 - 1.000] | 0.017 |

**Additional file 4.** Multivariate logistic regression analysis comparing wards patients against ICU patients (backward stepwise selection method / Likelihood Ratio). The association between viral RNAemia and viral RNA load with critical illness was evaluated adjusting by major confounding factors.

|  | OR [CI95%] |  |  | p | OR [CI95%] |  |  | p | OR [CI95%] |  |  | p |
| --- | --- | --- | --- | --- | --- | --- | --- | --- | --- | --- | --- | --- |
| Systolic Pressure (mmHg) | 0.976 | 0.953 | 1.000 | 0.049 | 0.974 | 0.949 | 0.999 | 0.040 | 0.974 | 0.950 | 0.999 | 0.042 |
| Glucose (mg/dl) | 1.006 | 1.000 | 1.012 | 0.050 | 1.006 | 1.000 | 1.012 | 0.055 | 1.005 | 0.999 | 1.012 | 0.088 |
| D-dimer (pg/mL) | 1.000 | 1.000 | 1.000 | 0.057 | 1.000 | 1.000 | 1.000 | 0.025 | 1.000 | 1.000 | 1.000 | 0.088 |
| LDH (UI/L) | 1.008 | 1.004 | 1.011 | 0.000 | 1.006 | 1.003 | 1.010 | 0.001 | 1.007 | 1.004 | 1.011 | 0.000 |
| Haematocrit (%) | 0.798 | 0.717 | 0.888 | 0.000 | 0.789 | 0.706 | 0.881 | 0.000 | 0.775 | 0.688 | 0.873 | 0.000 |
| Lymphocytes (cells/mm3) | 0.997 | 0.996 | 0.999 | 0.000 | 0.997 | 0.996 | 0.999 | 0.000 | 0.997 | 0.996 | 0.999 | 0.000 |
| Viral RNAemia (Yes) | 4.270 | 1.575 | 11.580 | 0.004 |  |  |  |  |  |  |  |  |
| Viral RNA load (N1)<br>in plasma, log (copies/mL) |  |  |  |  | 2.005 | 1.350 | 2.978 | 0.001 |  |  |  |  |
| Viral RNA load (N2)<br>in plasma, log (copies/mL) |  |  |  |  |  |  |  |  | 2.071 | 1.419 | 3.022 | 0.000 |

**Additional file 5.** Laboratory parameters ‘levels across groups.

|  | Healthy Controls<br>(0) | Outpatients<br>(1) | Ward<br>(2) | ICU<br>(3) | *p*<br>(0 vs 1) | *p*<br>(0 vs 2) | *p*<br>(0 vs 3) | *p*<br>(1 vs 2) | *p*<br>(1 vs 3) | *p*<br>(2 vs 3) |
| --- | --- | --- | --- | --- | --- | --- | --- | --- | --- | --- |
| ICAM-1<br>(pg/ml) | 289208<br>[92857] | 358431.50<br>[88702.30] | 377655<br>[150081.80] | 466818<br>[202189.80] | 0.036 | < 0.001 | < 0.001 | n.s. | < 0.001 | < 0.001 |
| Lipocalin-2<br>(pg/ml) | 82634.50<br>[29205.30] | 73134.50<br>[32007.30] | 94077<br>[49868] | 100165<br>[57819.80] | n.s. | n.s. | n.s. | 0.011 | < 0.001 | n.s. |
| Myeloperoxidase<br>(pg/ml) | 27541.50<br>[12906.80] | 41902<br>[30413.80] | 88244.50<br>[78145] | 141971.50<br>[148623.50] | n.s. | < 0.001 | < 0.001 | < 0.001 | < 0.001 | < 0.001 |
| VCAM-1<br>(pg/ml) | 618380<br>[195510.50] | 800393<br>[403378] | 1191387<br>[618827] | 1248699.50<br>[603679] | 0.021 | < 0.001 | < 0.001 | < 0.001 | < 0.001 | n.s. |
| PD-L1<br>(pg/ml) | 55.85<br>[33.27] | 110.50<br>[120.68] | 178<br>[114.25] | 228<br>[137.75] | 0.031 | < 0.001 | < 0.001 | 0.045 | < 0.001 | 0.031 |
| G-CSF<br>(pg/ml) | 6.90<br>[7.67] | 23.55<br>[20.48] | 31.95<br>[42.85] | 36.25<br>[53.35] | < 0.001 | < 0.001 | < 0.001 | n.s. | 0.022 | n.s. |
| IL-1b<br>(pg/ml) | 0.44<br>[1.65] | 0.16<br>[0.30] | 0.44<br>[0.53] | 0.38<br>[0.51] | n.s. | n.s. | n.s. | 0.005 | < 0.001 | n.s. |
| IL-10<br>(pg/ml) | 1.59<br>[0.48] | 4.19<br>[6.68] | 9.11<br>[10.14] | 17<br>[22.25] | 0.002 | < 0.001 | < 0.001 | 0.007 | < 0.001 | < 0.001 |
| IL-17A<br>(pg/ml) | 1.95<br>[1.08] | 2.38<br>[1.45] | 2.32<br>[1.44] | 3.06<br>[2.87] | n.s. | 0.047 | 0.002 | n.s. | n.s. | n.s. |
| GM-CSF<br>(pg/ml) | 0.66<br>[0.42] | 1.07<br>[0.54] | 1.10<br>[0.67] | 1.62<br>[1.01] | n.s. | n.s. | < 0.001 | n.s. | < 0.001 | < 0.001 |
| IL-7<br>(pg/ml) | 1.81<br>[1.49] | 2.04<br>[3.12] | 3.19<br>[4.19] | 4.84<br>[5.17] | n.s. | 0.009 | < 0.001 | 0.017 | < 0.001 | n.s. |
| CXCL10<br>(pg/ml) | 91.9<br>[71.85] | 470.50<br>[731.50] | 977.50<br>[943] | 1609.50<br>[1693.75] | 0.003 | < 0.001 | < 0.001 | 0.002 | < 0.001 | 0.003 |
| Angiopoietin-2<br>(pg/ml) | 708<br>[260.75] | 809<br>[479.75] | 1128<br>[779.50] | 1769.50<br>[2064.25] | n.s. | 0.001 | < 0.001 | 0.008 | < 0.001 | 0.001 |
| IL-1ra<br>(pg/ml) | 478<br>[1159.75] | 653.50<br>[1039.25] | 1796<br>[1896] | 1634<br>[4472.50] | n.s. | < 0.001 | < 0.001 | < 0.001 | < 0.001 | n.s |
| IL-6<br>(pg/ml) | 1.27<br>[0.83] | 6.34<br>[12.42] | 33.25<br>[48.10] | 105.50<br>[252.23] | 0.027 | < 0.001 | < 0.001 | < 0.001 | < 0.001 | < 0.001 |
| CCL2<br>(pg/ml) | 103.50<br>[39] | 189<br>[157.50] | 283.50<br>[304] | 490<br>[881.75] | 0.012 | < 0.001 | < 0.001 | 0.031 | < 0.001 | < 0.001 |
| IL-12p70<br>(pg/ml) | 0.62<br>[0.71] | 1.46<br>[0.67] | 1.51<br>[0.71] | 1.36<br>[0.94] | < 0.001 | < 0.001 | < 0.001 | n.s. | n.s. | n.s. |
| IL-2<br>(pg/ml) | 0.02<br>[0.09] | 0.53<br>[0.39] | 0.49<br>[0.57] | 0.63<br>[0.83] | < 0.001 | < 0.001 | < 0.001 | n.s. | n.s. | n.s. |
| IL-4<br>(pg/ml) | 0.13<br>[0.27] | 0.33<br>[0.34] | 0.33<br>[0.39] | 0.29<br>[0.37] | 0.021 | 0.006 | 0.041 | n.s. | n.s. | n.s. |
| IL-15<br>(pg/ml) | 2.11<br>[0.54] | 3.85<br>[3.7] | 4.89<br>[2.90] | 8.38<br>[5.19] | 0.001 | < 0.001 | < 0.001 | n.s. | < 0.001 | < 0.001 |
| Granzyme-B<br>(pg/ml) | 12<br>[15.75] | 33.20<br>[27.92] | 46.15<br>[40.03] | 39.25<br>[35.58] | < 0.001 | < 0.001 | < 0.001 | n.s. | n.s. | 0.036 |
| INFγ<br>(pg/ml) | 0.68<br>[0.45] | 4.35<br>[14.16] | 7.42<br>[15.13] | 3.44<br>[8.50] | < 0.001 | < 0.001 | < 0.001 | n.s. | n.s. | 0.021 |
| TNFα<br>(pg/ml) | 5.11<br>[1.79] | 10.60<br>[4.18] | 13<br>[6.46] | 12.95<br>[10.34] | < 0.001 | < 0.001 | < 0.001 | 0.021 | 0.006 | n.s. |

## References

1. COVID-19 Dashboard by the Center for Systems Science and Engineering (CSSE) at Johns Hopkins University (JHU) [Internet]. Available from: https://coronavirus.jhu.edu/map.html

2. Guan W-J, Ni Z-Y, Hu Y, Liang W-H, Ou C-Q, He J-X, et al. Clinical Characteristics of Coronavirus Disease 2019 in China. N Engl J Med. 2020;

3. Cevik M, Bamford CGG, Ho A. COVID-19 pandemic—a focused review for clinicians. Clinical Microbiology and Infection. Elsevier; 2020;26:842–7.

4. Zhang W, Du R-H, Li B, Zheng X-S, Yang X-L, Hu B, et al. Molecular and serological investigation of 2019-nCoV infected patients: implication of multiple shedding routes. Emerging Microbes & Infections. Taylor & Francis; 2020;9:386–9.

5. Chen X, Zhao B, Qu Y, Chen Y, Xiong J, Feng Y, et al. Detectable Serum Severe Acute Respiratory Syndrome Coronavirus 2 Viral Load (RNAemia) Is Closely Correlated With Drastically Elevated Interleukin 6 Level in Critically Ill Patients With Coronavirus Disease 2019. Clin Infect Dis [Internet]. [cited 2020 Jul 6]; Available from: https://academic.oup.com/cid/article/doi/10.1093/cid/ciaa449/5821311

6. Zheng S, Fan J, Yu F, Feng B, Lou B, Zou Q, et al. Viral load dynamics and disease severity in patients infected with SARS-CoV-2 in Zhejiang province, China, January-March 2020: retrospective cohort study. BMJ. 2020;369:m1443.

7. Varga Z, Flammer AJ, Steiger P, Haberecker M, Andermatt R, Zinkernagel AS, et al. Endothelial cell infection and endotheliitis in COVID-19. Lancet. 2020;395:1417–8.

8. Li H, Liu L, Zhang D, Xu J, Dai H, Tang N, et al. SARS-CoV-2 and viral sepsis: observations and hypotheses. Lancet. 2020;395:1517–20.

9. Leisman DE, Ronner L, Pinotti R, Taylor MD, Sinha P, Calfee CS, et al. Cytokine elevation in severe and critical COVID-19: a rapid systematic review, meta-analysis, and comparison with other inflammatory syndromes. The Lancet Respiratory Medicine [Internet]. Elsevier; 2020 [cited 2020 Oct 21];0. Available from: https://www.thelancet.com/journals/lanres/article/PIIS2213-2600(20)30404-5/abstract

10. Laing AG, Lorenc A, Del Molino Del Barrio I, Das A, Fish M, Monin L, et al. A dynamic COVID-19 immune signature includes associations with poor prognosis. Nat Med. 2020;

11. Bao J, Li C, Zhang K, Kang H, Chen W, Gu B. Comparative analysis of laboratory indexes of severe and non-severe patients infected with COVID-19. Clin Chim Acta. 2020;509:180–94.

12. Hupf J, Mustroph J, Hanses F, Evert K, Maier LS, Jungbauer CG. RNA-expression of adrenomedullin is increased in patients with severe COVID-19. Crit Care. 2020;24:527.

13. McGonagle D, O’Donnell JS, Sharif K, Emery P, Bridgewood C. Immune mechanisms of pulmonary intravascular coagulopathy in COVID-19 pneumonia. Lancet Rheumatol. 2020;2:e437–45.

14. Yang L, Liu S, Liu J, Zhang Z, Wan X, Huang B, et al. COVID-19: immunopathogenesis and Immunotherapeutics. Signal Transduct Target Ther. 2020;5:128.

15. Hogan CA, Stevens BA, Sahoo MK, Huang C, Garamani N, Gombar S, et al. High Frequency of SARS-CoV-2 RNAemia and Association With Severe Disease. Clin Infect Dis. 2020;

16. Veyer D, Kernéis S, Poulet G, Wack M, Robillard N, Taly V, et al. Highly sensitive quantification of plasma SARS-CoV-2 RNA shelds light on its potential clinical value. Clin Infect Dis. 2020;

17. Vabret N, Britton GJ, Gruber C, Hegde S, Kim J, Kuksin M, et al. Immunology of COVID-19: Current State of the Science. Immunity. 2020;52:910–41.

18. Cameron MJ, Ran L, Xu L, Danesh A, Bermejo-Martin JF, Cameron CM, et al. Interferon-mediated immunopathological events are associated with atypical innate and adaptive immune responses in patients with severe acute respiratory syndrome. J Virol. 2007;81:8692–706.

19. Cameron MJ, Bermejo-Martin JF, Danesh A, Muller MP, Kelvin DJ. Human immunopathogenesis of severe acute respiratory syndrome (SARS). Virus Res. 2008;133:13–9.

20. Bermejo-Martin JF, Ortiz de Lejarazu R, Pumarola T, Rello J, Almansa R, Ramírez P, et al. Th1 and Th17 hypercytokinemia as early host response signature in severe pandemic influenza. Crit Care. 2009;13:R201.

21. Rubio I, Osuchowski MF, Shankar-Hari M, Skirecki T, Winkler MS, Lachmann G, et al. Current gaps in sepsis immunology: new opportunities for translational research. Lancet Infect Dis. 2019;19:e422–36.

22. CDC. Information for Laboratories about Coronavirus (COVID-19) [Internet]. Centers for Disease Control and Prevention. 2020 [cited 2020 Oct 14]. Available from: https://www.cdc.gov/coronavirus/2019-ncov/lab/rt-pcr-panel-primer-probes.html

23. Hagman K, Hedenstierna M, Gille-Johnson P, Hammas B, Grabbe M, Dillner J, et al. SARS-CoV-2 RNA in serum as predictor of severe outcome in COVID-19: a retrospective cohort study. Clin Infect Dis. 2020;

24. de Jong MD, Simmons CP, Thanh TT, Hien VM, Smith GJD, Chau TNB, et al. Fatal outcome of human influenza A (H5N1) is associated with high viral load and hypercytokinemia. Nat Med. 2006;12:1203–7.

25. Almansa R, Anton A, Ramirez P, Martin-Loeches I, Banner D, Pumarola T, et al. Direct association between pharyngeal viral secretion and host cytokine response in severe pandemic influenza. BMC Infect Dis. 2011;11:232.

26. Birra D, Benucci M, Landolfi L, Merchionda A, Loi G, Amato P, et al. COVID 19: a clue from innate immunity. Immunol Res. 2020;68:161–8.

27. Perera P-Y, Lichy JH, Waldmann TA, Perera LP. The role of interleukin-15 in inflammation and immune responses to infection: implications for its therapeutic use. Microbes Infect. 2012;14:247–61.

28. Sugamata R, Dobashi H, Nagao T, Yamamoto K-I, Nakajima N, Sato Y, et al. Contribution of neutrophil-derived myeloperoxidase in the early phase of fulminant acute respiratory distress syndrome induced by influenza virus infection. Microbiol Immunol. 2012;56:171–82.

29. Venet F, Monneret G. Advances in the understanding and treatment of sepsis-induced immunosuppression. Nat Rev Nephrol. 2018;14:121–37.

30. Remy KE, Brakenridge SC, Francois B, Daix T, Deutschman CS, Monneret G, et al. Immunotherapies for COVID-19: lessons learned from sepsis. Lancet Respir Med. 2020;

31. Bermejo-Martin JF, Almansa R, Menendez R, Mendez R, Kelvin DJ, Torres A. Lymphopenic community acquired pneumonia as signature of severe COVID-19. Journal of Infection. 2020;

32. Monneret G, Cour M, Viel S, Venet F, Argaud L. Coronavirus disease 2019 as a particular sepsis: a 2-week follow-up of standard immunological parameters in critically ill patients. Intensive Care Med. 2020;46:1764–5.

